# Medication administration errors during general anesthesia – a systematic review of prospective studies

**DOI:** 10.1101/2023.03.28.23287875

**Authors:** Bradley P Murphy, Gayatri Sivaratnam, Jean Wong, Frances Chung, Amir Abrishami

**Author notes:** Bradley Murphy, (BPM).

## Abstract

**Introduction:** The incidence of medication error in anesthesia can be variable among different studies likely due to recall bias in retrospective studies. In prospective survey studies, questionnaires are sent to anesthesia care providers to facilitate self-reports of medication errors during a pre-planned follow-up period. This systematic review investigates all prospective survey studies of medication errors in adult patients undergoing general anesthesia. Our objective is to identify the incidence and characteristics of the common medication errors during general anesthesia. We also want to determine the contributing factors and outcomes of these errors.

**Methods:** We conducted database searches of Embase and Medline for medication errors in anesthesia between 1980 to 2019 and 2020 to 2021. Ten prospective survey studies detailing medication errors involving adult patients under general anesthesia were included. Data on response rate, incidence of errors, types of error and medications, patient outcomes, and contributing factors were collected.

**Results:** Ten studies were included of which six studies provided a response rate ranging from 53% to 97.5%. The incidence of medication errors ranged from 0.02% to 1.12% or 1 in every 90 to 5000 anesthetics. A total of 1,676 medication errors during general anesthesia were analyzed. The most reported error was the substitution error (31.6% [530/1676]), followed by incorrect dose (28.4% [476/1676]). The class of medication most associated with administration errors were muscle relaxants, opioids, and antibiotics. Most patient outcomes were of no harm. Inexperience of the anesthesiologist, nurse or student was the most reported contributing factor, followed by haste or pressure to proceed, and communication problems.

**Conclusion:** The incidence of medication errors during general anesthesia were as high as 1.12% and the most common errors were substitution error and incorrect dose. Inexperience, time pressure, and communication problems were contributing factors. This information can be used to inform safety practices in anesthesia.

## 1 Introduction

During administration of anesthesia, multiple drugs are given from different classes via various routes in fast-paced and high intensity environments.(1) Given the nature of anesthesia, in which most anesthesiologists administer over 250,000 drugs during their career,(2) medication errors are of significant concern. Anesthesiologists are likely to make one or more errors or near errors over the span of their careers.(3–5)

Cognizant of this increased potential for harm, anesthesia is a leading specialty in patient safety. The error rate in anesthesia has decreased over the last few decades, such that operative mortality due to anesthesia is 1 per 100,000.(6) Many studies have shown medication errors to be one of the main causes of adverse events in anesthesia.(7–17) These errors cause iatrogenic harm and increase healthcare costs through increased length of stay (LOS) and surgery times.(4) Recognizing these implications, a recent study highlights the work to develop and disseminate consensus-based recommendations for perioperative medication safety.(18) Of note, many of the existing recommendations that aim to prevent perioperative medication errors are not feasible in middle-income and low-income countries.(18) Thus, the recommendations are tailored to income level of each country.

Medication errors refer to situations in which a drug is erroneously administered and can occur at any point throughout the anesthetic process.(1) While the many causes and types of medication errors within anesthesia have been investigated, there is considerable variation among studies, leading to uncertainty regarding its magnitude.(19) There are studies with different designs such as case report/series, databases reviews, and retrospective or prospective survey studies that investigated the occurrence and outcomes of medication errors in anesthesia. These studies have been mostly conducted at an institutional or national level. In survey studies, questionnaires are sent to anesthesia care providers to facilitate self-reports of any medication errors that occurred in the past (retrospective recall) or during a follow up period (prospective survey).

To date, there have been minimal efforts to summarize the current evidence on this topic likely due to complexity and abundancy of the literature. A recent integrated review by Bratch et al.,(20) included a variety of study designs to analyze the incidence and type of medication errors during anesthesia. The combination of diverse methodologies can lead to inaccurate results and difficulties in drawing conclusions in reviews.(21) Including retrospective studies within their report raises concern for recall and reporting bias. Although there may be an overestimate with prospective studies, this emphasizes the need for forming a standardized definition of medication errors to properly capture this data in future studies.

As per the 2017 WHO Medication Without Harm patient safety initiative, their vision was to reduce the level of severe, avoidable harm related to medications by 50% over 5 years.(22) However, given the heterogeneity between studies, the true rate of medication errors in anesthesia is not fully known. The objectives of our systematic review are to gather data from prospective studies to investigate the response rate, incidence of errors, common medication errors, implicated medications, outcomes of medications errors, and their contributing factors. Determining the most common causes of medication errors reveal error-prone practices, allowing us to develop strategies to avoid mistakes, ultimately improving safety of anesthetic practice.

## 2 Methods

### 2.1 Protocol

This systematic review was created and conducted in accordance with the Preferred Reporting Items for Systematic Review and Meta-Analyses (PRISMA) guidelines.

### 2.2 Search strategy

The Embase and Medline databases were searched using a search strategy in collaboration with our librarian at McMaster University. The keywords were “anesthesia”, “anesthetic agent” and “medication errors”, “patient safety” and its related keywords. The citation lists of included articles were thoroughly reviewed to capture any articles that were potentially missed from the original search. The search was limited to English language and humans. The search strategy is attached in S1 Table.

### 2.3 Study selection and data extraction

The search was conducted from January 1, 1980, to December 31, 2019 by authors GS and AA, with an updated search from January 1, 2020 to December 11, 2021 completed by authors BM and AA. Continued surveillance of literature was done up to November 2022. After duplicates were removed, the title, abstracts, and full text of the eligible studies were reviewed in a stepwise fashion and irrelevant studies were excluded.

Studies were included in this review if they met the following criteria: 1) All prospective studies on medication errors related to the anesthetic process, 2) patients aged 18 years and older having surgery under general anesthesia, and 3) publications in English. We excluded case reports, case series, quality improvement studies, systematic reviews, meta-analyses, review articles, database reviews, retrospective surveys, cohort studies, and cross-sectional studies.

### 2.4 Data analysis

Data on the response rate and incidence of errors, error and near miss frequency, types of error, involved medications, patient outcomes, and contributing factors were collected. Due to the heterogeneity in patient outcomes across studies, the following categorization system was used to allow uniformity in data collection and presentation across studies.(23) No harm refers to an error that did not cause harm; error resulted in the need for additional monitoring or tests but no harm. Mild harm refers to a harmful effect that was mild, temporary, and short-term; no treatment or only minor treatment was required. Moderate harm refers to a harmful effect that required more than minor treatment (including procedural treatment) or required an unplanned hospital admission or prolonged hospital stay. Severe harm refers to symptoms that required major treatment to save the patient’s life or caused major permanent or long-term harm.

Quality assessment was not done as there were no critical appraisal tools available to assess the quality of the prospective survey studies.

## 3 Results

### 3.1 Search strategy

The literature search yielded 15,998 citations (Fig 1). After we removed duplicates, 14,262 studies remained. After we screened titles and abstracts, we found 72 articles to be eligible for full-text review. Of these articles, three studies met the inclusion criteria. We identified an additional seven records through the citation search (Table 1). A total of ten studies were included for final review.(2,23–31)

**Fig 1.**
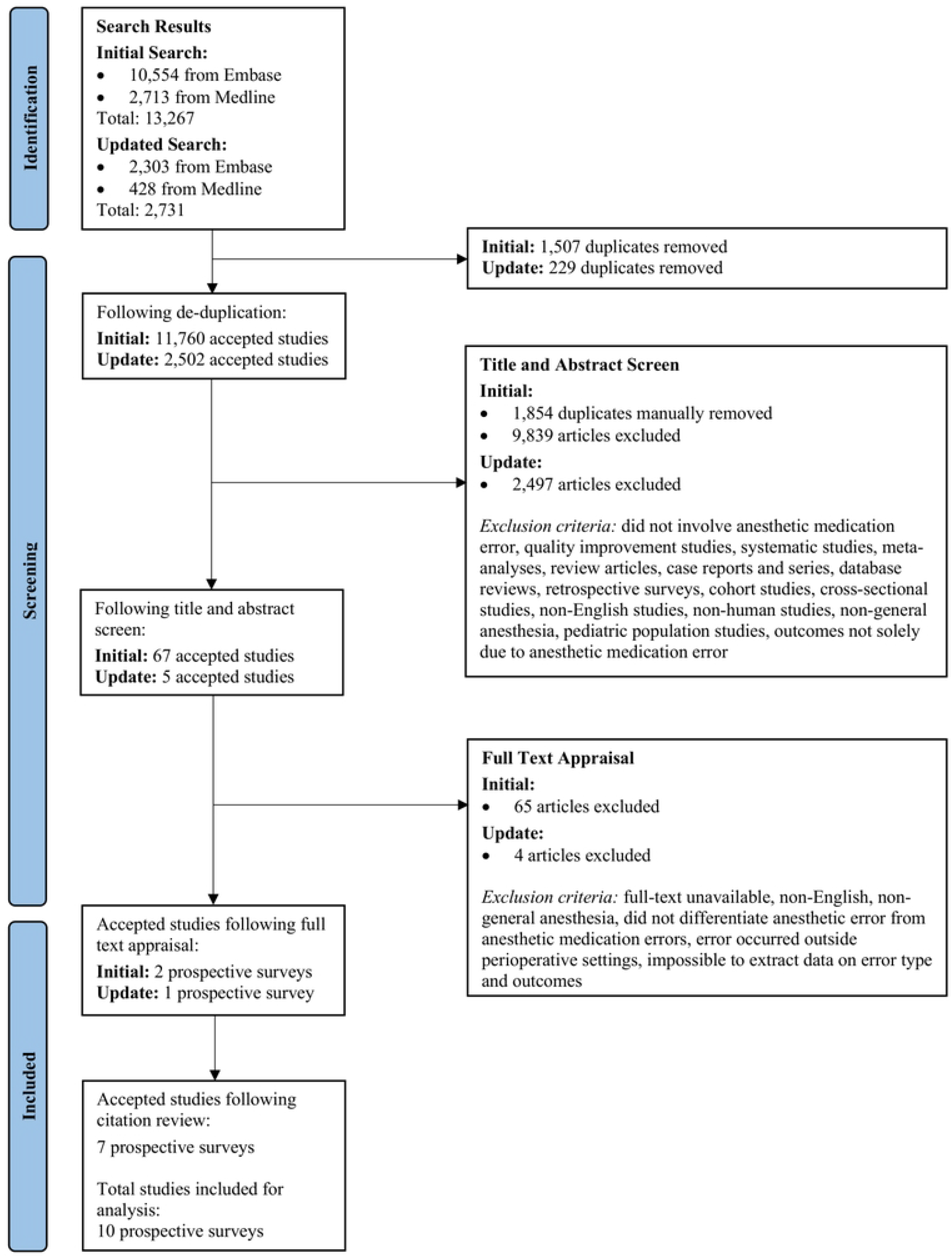
Search protocol for including and excluding studies within this systematic review.

**Table 1.** Characteristics of the ten prospective surveys.

| Citation | Country | Study Duration | Study Population | Practice Setting | Total Number of Anesthetics | Response Rate (%) | Incidence of Errors (%) |
| --- | --- | --- | --- | --- | --- | --- | --- |
| Kim 2022 | Australia, New Zealand | 6 Years | Physicians | Hospitals (Not specified) | 4,000* | - | - |
| Bowdle 2018 | USA | 14 Years | Physicians, nurses | University hospital | 51,846 | 97.5 | 0.44 |
| Zhang 2013 | China | 6 Months | Physicians | Tertiary hospital | 24,380 | 67.7 | 0.73 |
| Cooper 2012 | USA | 6 Months | Physicians, nurses | Tertiary teaching hospital | 10,574 | 83 | 0.49 |
| Webster 2010 | New Zealand | 4-5 Years | Physicians | Tertiary teaching hospitals | 74,478 | 79.6 | 0.44 |
| Llewellyn 2009 | South Africa | 6 Months | Physicians | Tertiary teaching hospitals | 30,412 | 53 | 0.36 |
| Yamamoto 2008 | Japan | 8 Years | Physicians | University hospital | 27,454 | - | 0.17 |
| Hintong 2005 | Thailand | 18 Months | Physicians, nurses, students | University, tertiary, secondary, and primary care hospitals | 202,699 | - | 0.02 |
| Webster<br>2001 | New<br>Zealand | 4-18<br>Months | Physicians | Tertiary<br>teaching<br>hospitals | 10,806 | 72.1 | 1.12 |
| Fasting<br>2000 | Norway | 3 Years | Physicians,<br>nurses | University<br>hospital | 55,426 | - | 0.11 |

\*Over 4000 incident reports were received in this study duration, however, this study only analyzed the first 4000 reports.

### 3.2 Study characteristics

Ten prospective studies published between 2000 to 2022 were included for the final review (Table 1). In these studies, anesthesia care providers were asked to complete and return a study form (i.e., medication error survey or incident forms) anonymously for every anesthetic performed during a set period. They were asked to indicate whether a drug administration error, or in some studies, a near miss (an incident with the potential to become an error) had occurred or not, and if the prior was affirmed, further details were elicited. These studies were conducted in different settings and multiple regions: New Zealand (n = 3), United States (n = 2), Australia (n = 1), China (n=1), Japan (n = 1), Norway (n = 1), South Africa (n = 1), and Thailand (n = 1). Of the ten studies, six provided a response rate which ranged from 53% to 97.5%. The incidence of medication errors ranged from 0.02% to 1.12% or 1 in every 90 to 5000 anesthetics.

### 3.3 Medication errors

A total of 1,676 anesthesia medication errors were analyzed, which included both errors and near misses (Table 2). Both errors and near misses were combined together when delineating the type of medication error. We found that the most common type of errors were substitution, incorrect dose, omission, incorrect route, repetition, insertion, and other (Table 3). Three studies listed these types of error and defined each type in their study form. (25,27,30) The most reported medication error in ten studies is the substitution error (31.6% [530/1676]), followed by incorrect dose (28.4% [476/1676]) (Table 3). The third most common error is the error of omission being reported in nine out of ten studies.

**Table 2.**
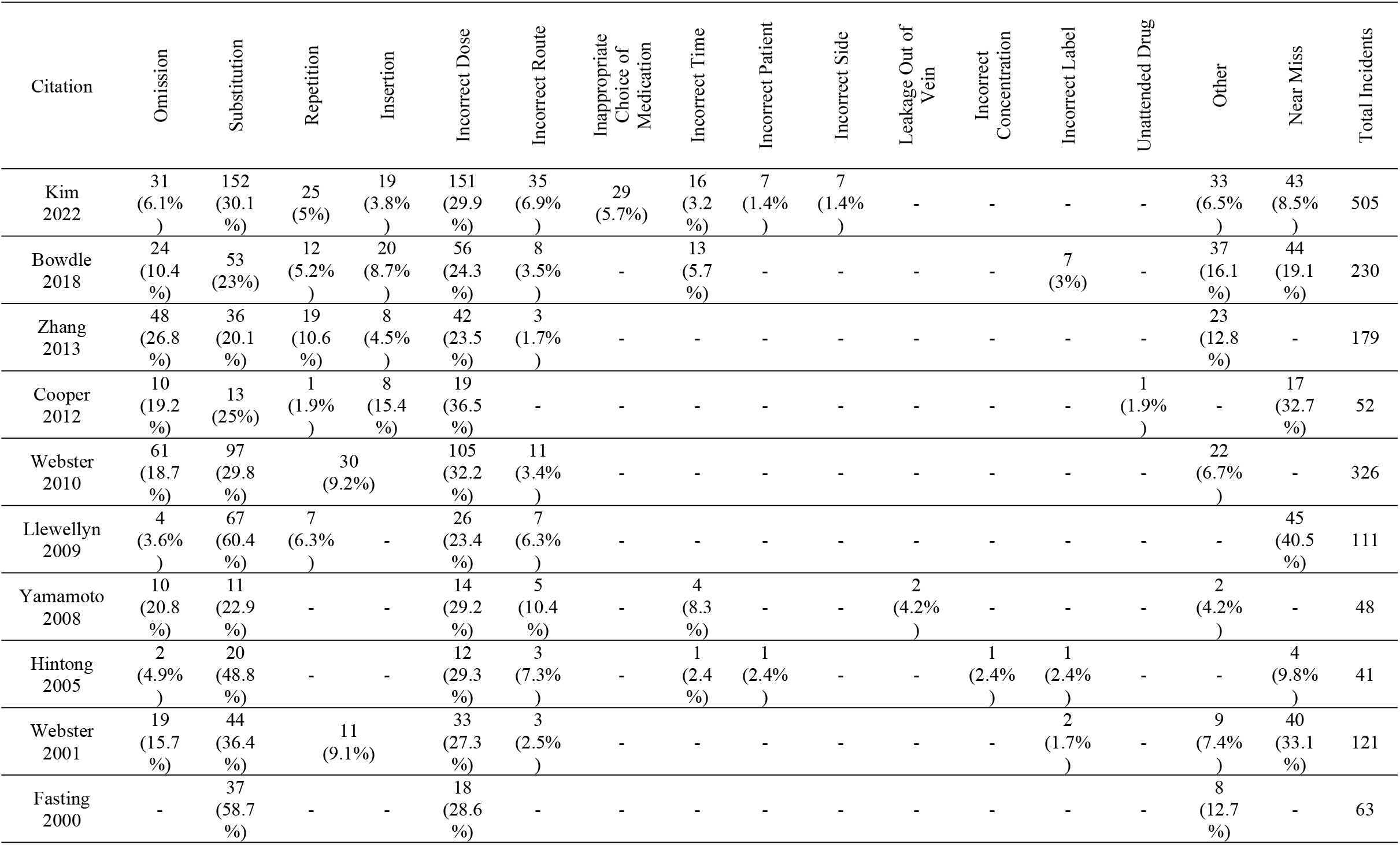
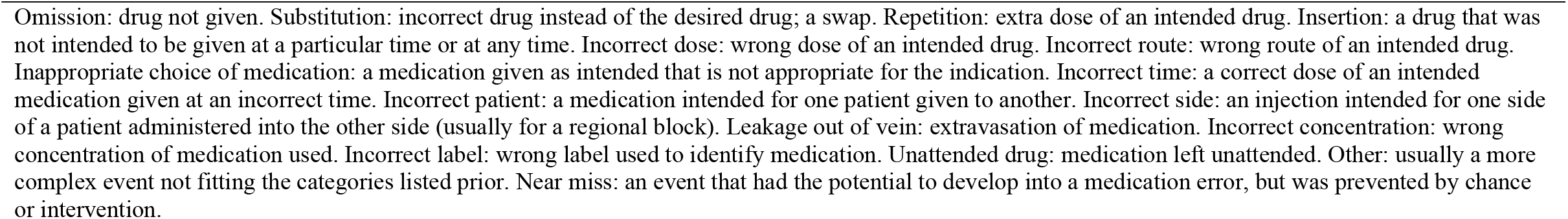
Reported medication errors.

**Table 3.**
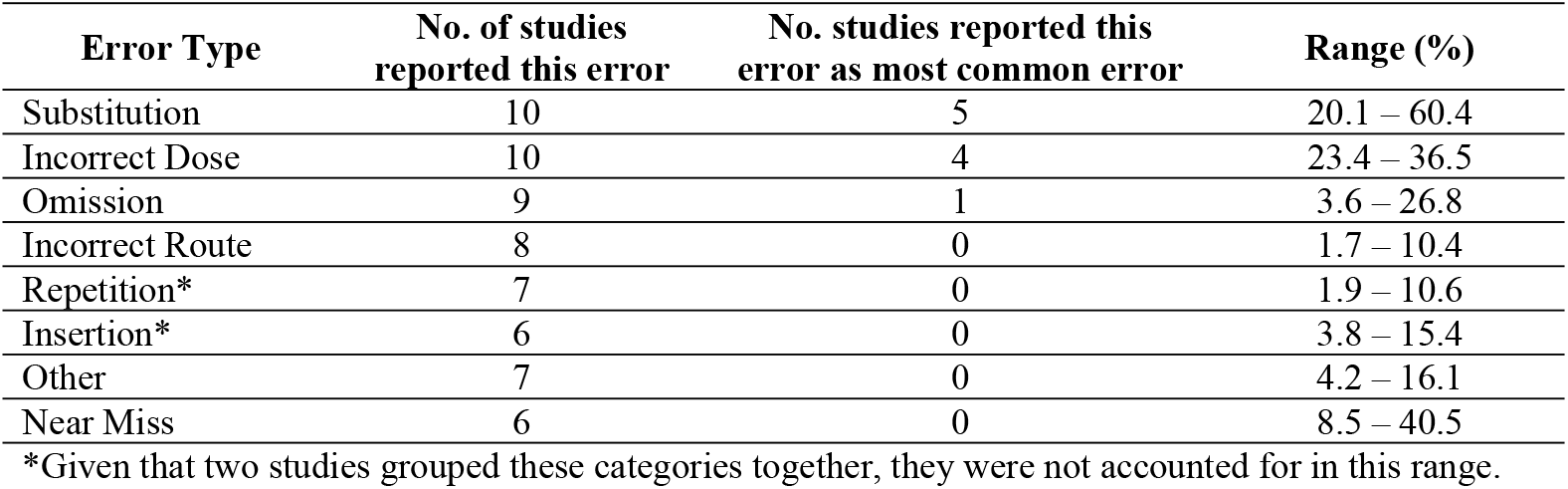
Ranges of most reported errors by error type.

| Error Type | No. of studies reported this error | No. studies reported this error as most common error | Range (%) |
| --- | --- | --- | --- |
| Substitution | 10 | 5 | 20.1 – 60.4 |
| Incorrect Dose | 10 | 4 | 23.4 – 36.5 |
| Omission | 9 | 1 | 3.6 – 26.8 |
| Incorrect Route | 8 | 0 | 1.7 – 10.4 |
| Repetition* | 7 | 0 | 1.9 – 10.6 |
| Insertion* | 6 | 0 | 3.8 – 15.4 |
| Other | 7 | 0 | 4.2 – 16.1 |
| Near Miss | 6 | 0 | 8.5 – 40.5 |

\*Given that two studies grouped these categories together, they were not accounted for in this range.

Of the ten studies, only seven reported the types of medication classes (S2 Table). The types of medication classes involved in errors varied. If a study listed only the medication names, these were categorized based on their medication class to present the data in a similar format. The medication class most associated with medication errors are muscle relaxants, opioids, and antibiotics (Fig 2). The incidence of errors related to the different classes of medication is as follows: (22.9% [8/35]), opioids (20% [7/35]), antibiotics (17.1% [6/35]), inhalational agents (11.4% [4/35]), local anesthetics (8.6% [3/35]), non-opioid analgesics (8.6% [3/35]), anticholinergics (5.7% [2/35]), induction agents (2.9% [1/35]), and sympathomimetics (2.9% [1/35]).

**Fig 2.**
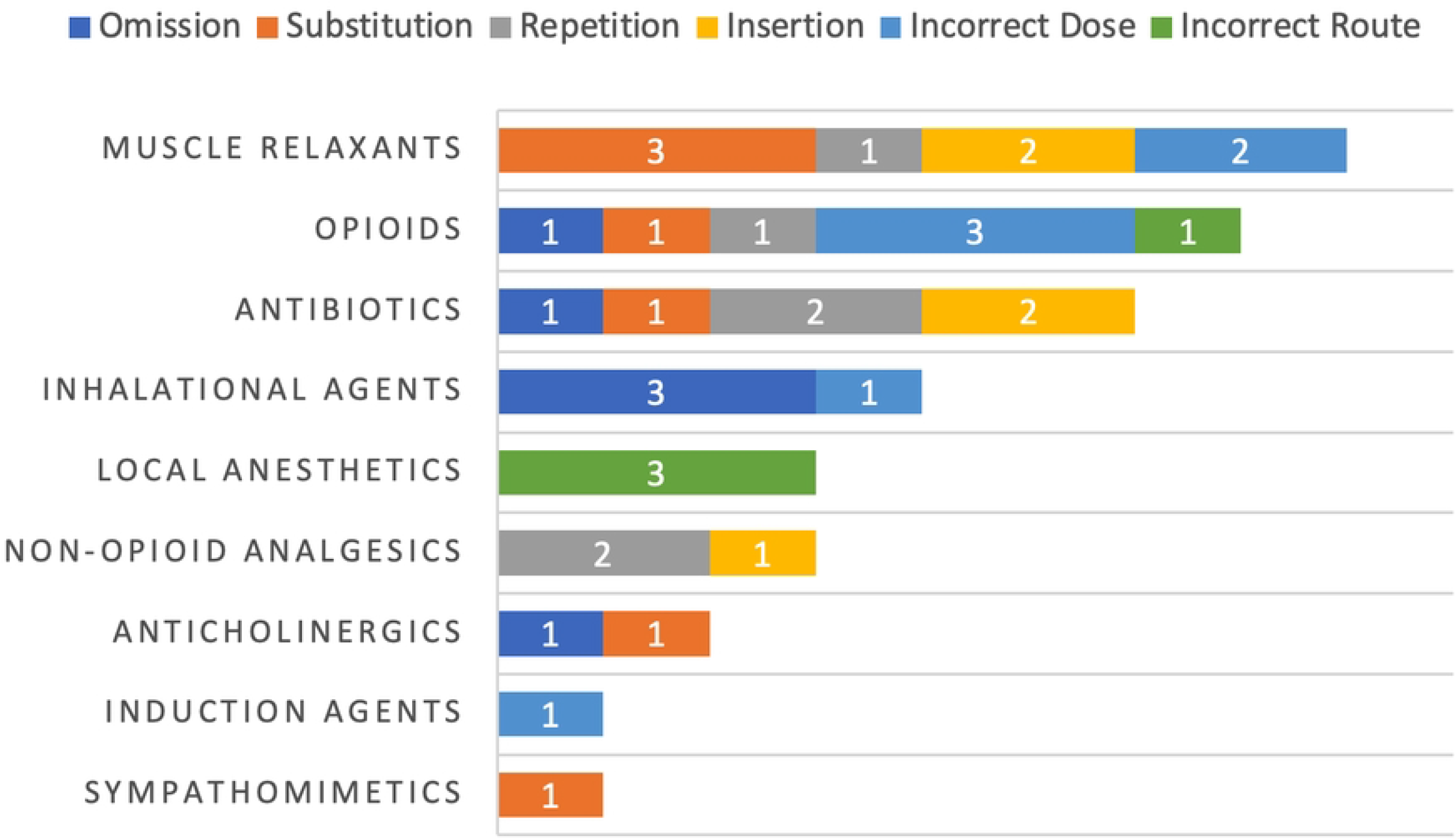
Most common type of medications involved in medication errors versus type of errors. Number of studies where medication class is listed as most common type of error is listed within each block.

When analyzing the most common types of error and medication classes (Fig 2), we found that substitution errors (20% [7/35]) and incorrect dosing errors (20% [7/35]) were the most common error type. Of the eight reported errors for muscle relaxants, three were substitution errors, two insertion errors, two incorrect dosing errors, and one a repetition error. Of the seven reported errors for opioids, three were incorrect dosing errors, one error each for omission, substitution, repetition, and incorrect route.

### 3.4 Patient outcomes

Patient outcomes that were contributed to medication errors ranged from no harm to severe harm (Table 4). The moderate and severe harm made up the minority of outcomes, ranging from 0.9% to 28.3%. Most errors resulted in no harm, ranging from 35% to 100%. Examples of severe harm reported included cardiac arrest, shock, respiratory depression, major morbidity, and death.

**Table 4.**
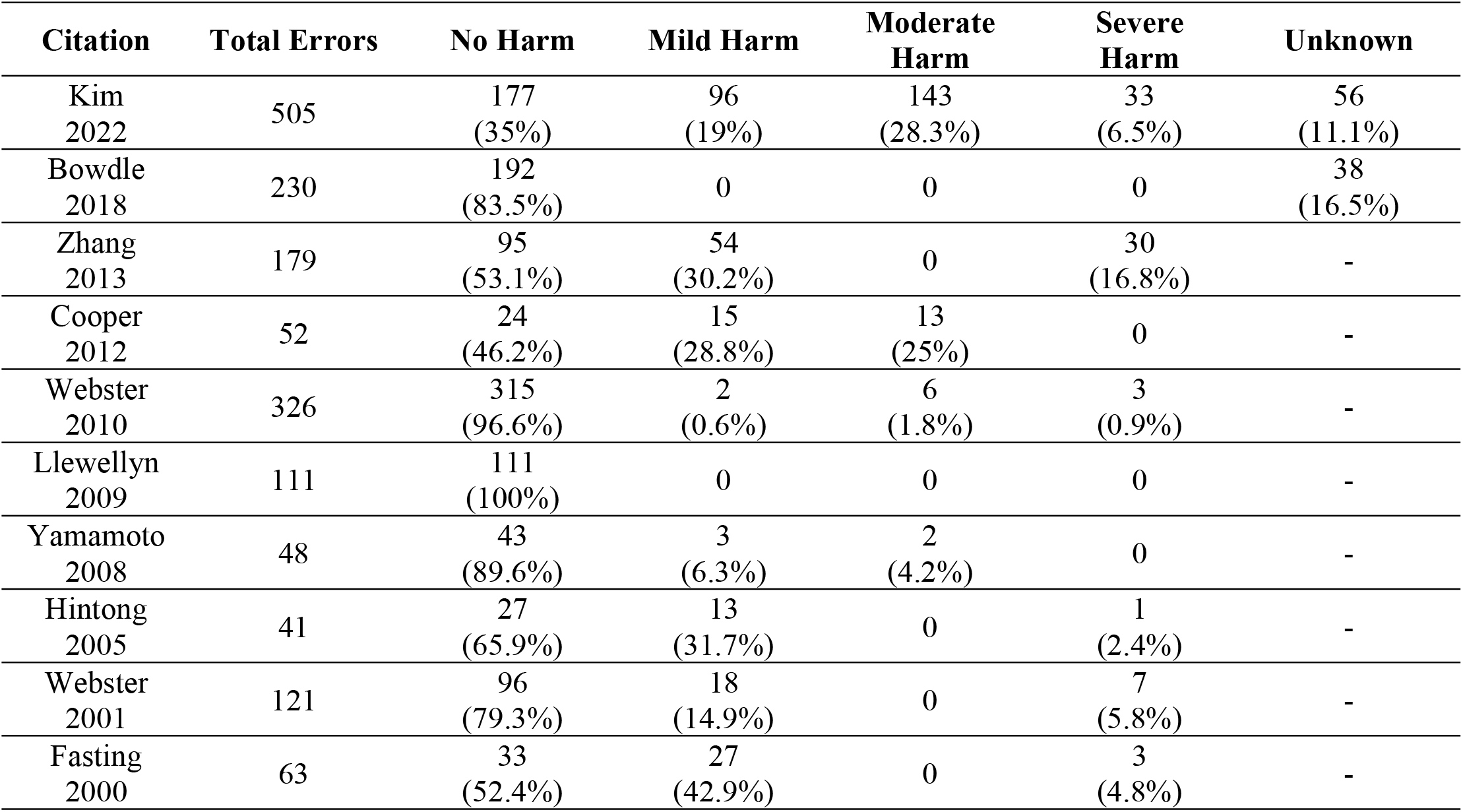
Outcomes of medication errors.

| Citation | Total Errors | No Harm | Mild Harm | Moderate Harm | Severe Harm | Unknown |
| --- | --- | --- | --- | --- | --- | --- |
| Kim 2022 | 505 | 177<br>(35%) | 96<br>(19%) | 143<br>(28.3%) | 33<br>(6.5%) | 56<br>(11.1%) |
| Bowdle 2018 | 230 | 192<br>(83.5%) | 0 | 0 | 0 | 38<br>(16.5%) |
| Zhang 2013 | 179 | 95<br>(53.1%) | 54<br>(30.2%) | 0 | 30<br>(16.8%) | - |
| Cooper 2012 | 52 | 24<br>(46.2%) | 15<br>(28.8%) | 13<br>(25%) | 0 | - |
| Webster 2010 | 326 | 315<br>(96.6%) | 2<br>(0.6%) | 6<br>(1.8%) | 3<br>(0.9%) | - |
| Llewellyn 2009 | 111 | 111<br>(100%) | 0 | 0 | 0 | - |
| Yamamoto 2008 | 48 | 43<br>(89.6%) | 3<br>(6.3%) | 2<br>(4.2%) | 0 | - |
| Hintong 2005 | 41 | 27<br>(65.9%) | 13<br>(31.7%) | 0 | 1<br>(2.4%) | - |
| Webster 2001 | 121 | 96<br>(79.3%) | 18<br>(14.9%) | 0 | 7<br>(5.8%) | - |
| Fasting 2000 | 63 | 33<br>(52.4%) | 27<br>(42.9%) | 0 | 3<br>(4.8%) | - |

### 3.5 Contributing factors

The contributing factors for these medication errors were listed in nine of the ten studies. S3 Table lists each study and their associated contributing factors. Each contributing factor was given a single point value and were totalled to determine which factors were most associated with a medication error (Fig 3). Inexperience was found to be the most reported factor, followed by haste or pressure to proceed and communication problems.

**Fig 3.**
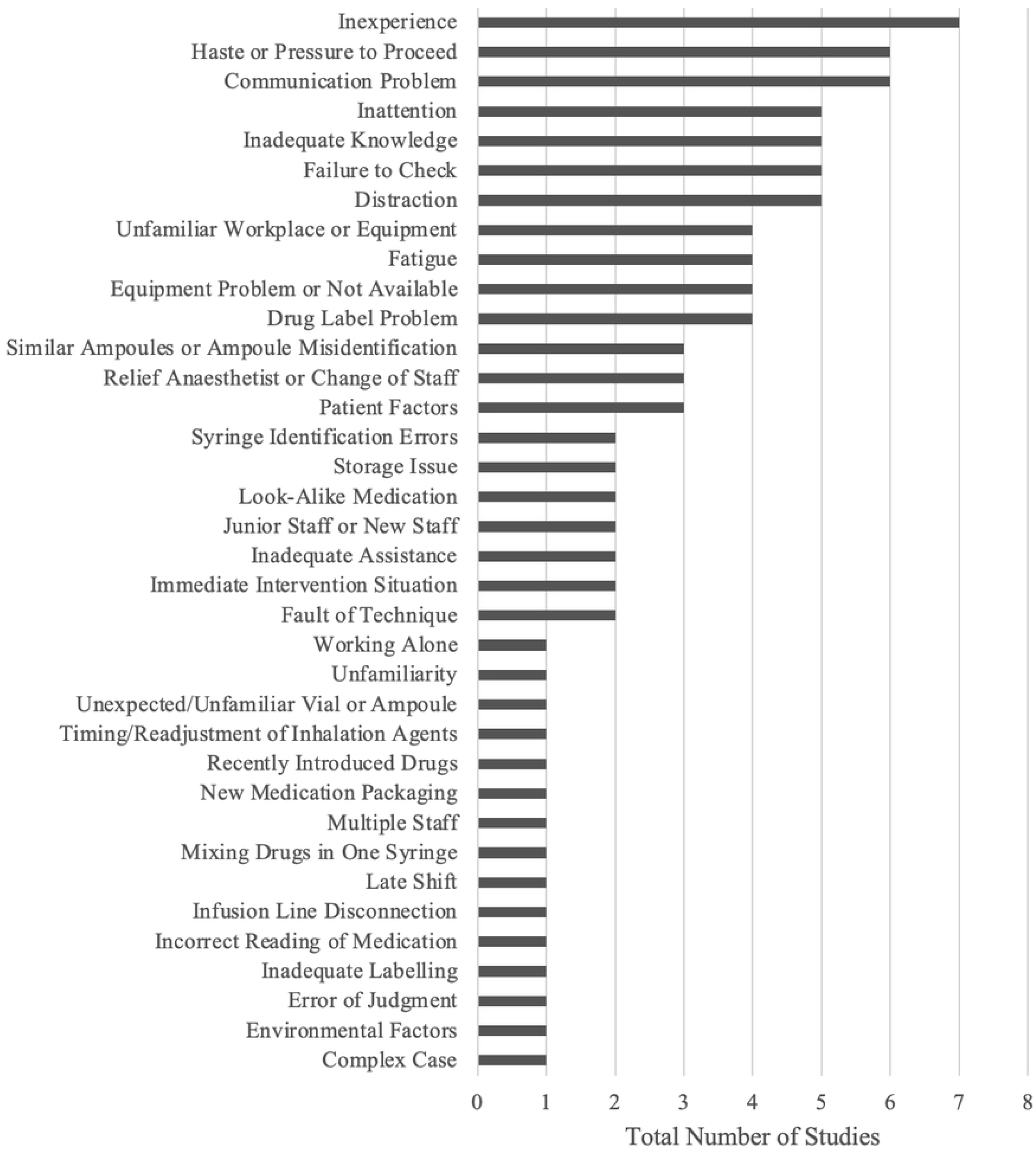
Contributing factors associated with medication errors.

## 4 Discussion

In this systematic review of ten prospective studies on medication errors in adults undergoing general anesthesia, we found that substitution errors and incorrect dosing errors were the most reported errors. The medication class most associated with administration errors were muscle relaxants, opioids, and antibiotics. Most patient outcomes were of no harm; however, several studies did report severe harm. Inexperience of the anesthesiologist, nurse or student was the most reported contributing factor, followed by haste or pressure to proceed, and communication problems.

Retrospective surveys and database reviews have previously identified substitution and incorrect dosing errors as the most commonly reported error type.(4,7,32–36) To date, the literature examining the involved medications, patient outcomes, and contributing factors of these medication errors are limited.(20)

Standardizing anesthetic procedures has often been suggested as a method of error reduction. The study completed by Maximous et al.(37) identified that multimodal interventions and improved labelling practices reduce medication errors in anesthetized patients. Multimodal interventions included a barcode reader that provided automatic auditory and visual verification of the drug selected, improved anesthetic carts, and other components in the bundle, such as colour-coded syringes, pre-filled syringes, reorganization of the workspace, and improved guidelines.(37) Of the six studies that reported using various combinations of these interventions into bundles, there was a significant reduction in the rate of errors post-intervention, ranging from 21% to 100%.(37) With respect to improved labelling systems, one study showed a 37% reduction in rate of errors, but this did not reach statistical significance.(37) Although multimodal interventions are becoming more common, globally many of these interventions, such as prefilled syringes or scanned labels, are not available for anesthesia practice. Additionally, many of these interventions come with a significant cost to implement.

We found that muscle relaxants, opioids, and antibiotics were most associated with medication errors during general anesthesia. This highlights an important finding, especially errors related to muscle relaxants. Errors with muscle relaxants can lead to important consequences, such as awareness, delayed recovery, or postoperative residual paralysis. While two-person checks for antibiotic administration is a common practice in many operating rooms, opioids and muscle relaxants are mainly administrated by anesthesia care providers. Therefore, other strategies such as barcode read outs at the time of administration could potentially reduce risk of errors.(24) Adding barcode labels to pre-filled syringes so computers can scan and “read out” labels, would provide the anesthesiologist information via audition, rather than simply reading the medication name.(16) While this heuristic can be timesaving, it can also be harmful when medications have similar names that can be misinterpreted by pattern recognition. Additionally, using barcodes would aid documentation of medication administration.(16) Although most studies reported that the medication errors resulted in no harm, several studies reported severe harm such as death, cardiac arrest, shock, respiratory depression, and major morbidity. This highlights the importance of continuing to improve anesthesia safety practices to minimize any potential for severe harm to patients undergoing general anesthesia.

In our systematic review, the most significant contributing factor was inexperience of the anesthesiologist, nurse, or student. Haste or pressure to proceed and communication problems were other contributing factors. Our findings are in contrary to those in retrospective surveys in which the most common contributing factor was syringe swap, followed by misidentification.(4,7,36) This highlights the potential recall bias that occurs when data is collected retrospectively in comparison to a prospective manner.

### Future direction

Future research should elucidate further which drugs are most often implicated in medication errors, as this could inform safety practices. Near errors should be elucidated to determine what prevented these incidents developed into completed errors. Importantly, the cause of errors, whether active or latent human error should be studied. The former would indicate changes to training of anesthesiologists, while the latter would inform system level changes within hospitals.

Future studies should utilize a standardized definition of medication errors, as this will allow for more comprehensive data analysis between studies and provide a more accurate representation on medication error rates.

### Strengths and limitations of study

Our study is the first to examine only prospective studies. In comparison to Bratch et al.,(20) where a variety of study designs were analyzed, we examined prospective surveys to minimize recall bias and provide an accurate picture of the true incidence of medication errors. There are some limitations in this review. Limitations stem from the large variance in data presentation within the studies. This large variability raises the need for standardized definitions of medications errors. Many studies did not link medications or medication class with error type. Also, some studies combined the medication errors and near misses together before delineating the type of error. Surveys rely on self-reporting, a biased modality of data collection. Additionally, anesthesia may only partially contribute to an incident, but may not be solely responsible. Near misses may not be recognized. Reluctance to report or inability to admit error can lead to underreporting.

## 5 Conclusion

Our systematic review found that anesthesiologists, as well as nurses and students, are most prone to errors of substitution. During general anesthesia, muscle relaxants, opioids, and antibiotics are the medication classes most associated with medication errors. Inexperience was the most reported contributing factor to medication errors. Using these information, new safety endeavors can be created to further ensure patient safety within the practice of anesthesia.

## Data Availability

All relevant data are within the manuscript and its Supporting Information files.

## Supporting information

**S1 Table. Search strategy for anesthetic medication errors in the Embase and Medline databases**.

**S2 Table. Medication classes involved by error type (medication class occurring at the highest frequency listed first)**.

**S3 Table. Contributing factors listed by study**.

## Notes

### Competing Interest Statement

The authors have declared no competing interest.

### Funding Statement

The author(s) received no specific funding for this work.

## References

1. Paix AD, Bullock MF, Runciman WB, Williamson JA. Crisis management during anaesthesia: problems associated with drug administration during anaesthesia. BMJ Qual Saf. 2005;14:e15.

2. Llewellyn RL, Gordon PC, Wheatcroft D, Lines D, Reed A, Butt AD, et al. Drug administration errors: a prospective survey from three South African teaching hospitals. Anaesth Intensive Care. 2009;37(1):93–8.

3. Merry AF, Peck DJ. Anaesthetists, errors in drug administration and the law. Vol. 108, New Zealand Medical Journal. 1995. p. 185–7.

4. Gordon PC, Llewellyn RL, James MFM. Drug administration errors by South African anaesthetists – a survey. South African Medical Journal. 2006;96(7):630–2.

5. Annie SJ, Thirilogasundary MR, Hemanth Kumar VR. Drug administration errors among anesthesiologists: The burden in India - A questionnaire-based survey. J Anaesthesiol Clin Pharmacol. 2019;35(2):220–6.

6. Mellin-Olsen J, Staender S, Whitaker DK, Smith AF. The Helsinki Declaration on Patient Safety in Anaesthesiology. Eur J Anaesthesiol. 2010;27(7):592–7.

7. Orser BA, Chen RJB, Yee DA. Medication errors in anesthetic practice: a survey of 687 practitioners. Canadian Journal of Anesthesia. 2001;48(2):139–46.

8. Nanji KC, Patel A, Shaikh S, Seger DL, Bates DW. Evaluation of Perioperative Medication Errors and Adverse Drug Events. Anesthesiology. 2016;124(1):25–34.

9. Glavin RJ. Drug errors: consequences, mechanisms, and avoidance. Vol. 105, British Journal of Anaesthesia. 2010. p. 76–82.

10. Kothari D, Gupta S, Sharma C, Kothari S. Medication error in anaesthesia and critical care: A cause for concern. Vol. 54, Indian Journal of Anaesthesia. 2010. p. 187–92.

11. Mahajan RP. Medication errors: Can we prevent them? Vol. 107, British Journal of Anaesthesia. 2011. p. 3–5.

12. Hove LD, Steinmetz J, Christoffersen JK, Møller A, Nielsen J, Schmidt H. Analysis of deaths related to anesthesia in the period 1996-2004 from closed claims registered by the Danish Patient Insurance Association. Anesthesiology. 2007;106(4):675–80.

13. Sakaguchi Y, Tokuda K, Yamaguchi K, Irita K. Incidence of anesthesia-related medication errors over a 15-year period in a university hospital. Fukuoka Igaku Zasshi. 2008;99(3):58–66.

14. Irita K, Tsuzaki K, Sawa T, Sanuki M, Makita K, Kobayashi Y, et al. Critical incidents due to drug administration error in the operating room: An analysis of 4,291,925 anesthetics over a 4 year period. Japanese Journal of Anesthesiology. 2004;53(5):577–84.

15. Cooper JB, Newbower RS, Kitz RJ. An analysis of major errors and equipment failures in anesthesia management: Considerations for prevention and detection. Anesthesiology. 1984;60(1):34–42.

16. Neily J, Silla ES, Sum-Ping Sjt, Reedy R, Paull DE, Mazzia L, et al. Anesthesia Adverse Events Voluntarily Reported in the Veterans Health Administration and Lessons Learned. Anesth Analg. 2018;126(2):471–7.

17. Kurth CD, Tyler D, Heitmiller E, Tosone SR, Martin L, Deshpande JK. National pediatric anesthesia safety quality improvement program in the United States. Anesth Analg. 2014;119(1):112–21.

18. Nanji KC, Merry AF, Shaikh SD, Pagel C, Deng H, Wahr JA, et al. Global PRoMiSe (Perioperative Recommendations for Medication Safety): protocol for a mixed-methods study. BMJ Open. 2020;10(6):e038313.

19. Nwasor EO, Sule ST, Mshelia DB. Audit of medication errors by anesthetists in North Western Nigeria. Niger J Clin Pract. 2014;17(2):226–31.

20. Bratch R, Pandit JJ. An integrative review of method types used in the study of medication error during anaesthesia: implications for estimating incidence. Br J Anaesth. 2021;127(3):458–69.

21. Hopia H, Latvala E, Liimatainen L. Reviewing the methodology of an integrative review. Scand J Caring Sci [Internet]. 2016 Dec 1;30(4):662–9. Available from: https://doi.org/10.1111/scs.12327

22. WHO, World Health Organization. Medication Without Harm. Medication Without Harm. 2022.

23. Kim JY, Moore MR, Culwick MD, Hannam JA, Webster CS, Merry AF. Analysis of medication errors during anaesthesia in the first 4000 incidents reported to webAIRS. Anaesth Intensive Care. 2022;50(3):204–19.

24. Bowdle TA, Jelacic S, Nair B, Togashi K, Caine K, Bussey L, et al. Facilitated self-reported anaesthetic medication errors before and after implementation of a safety bundle and barcode-based safety system. Br J Anaesth. 2018;121(6):1338–45.

25. Zhang Y, Dong YJ, Webster CS, Ding XD, Liu XY, Chen WM, et al. The frequency and nature of drug administration error during anaesthesia in a Chinese hospital. Acta Anaesthesiol Scand. 2013;57(2):158–64.

26. Cooper L, DiGiovanni N, Schultz L, Taylor AM, Nossaman B. Influences observed on incidence and reporting of medication errors in anesthesia. Canadian Journal of Anesthesia. 2012;59(6):562–70.

27. Webster CS, Larsson L, Frampton CM, Weller J, McKenzie A, Cumin D, et al. Clinical assessment of a new anaesthetic drug administration system: a prospective, controlled, longitudinal incident monitoring study. Anaesthesia. 2010;65(5):490–9.

28. Yamamoto M, Ishikawa S, Makita K. Medication errors in anesthesia: An 8-year retrospective analysis at an urban university hospital. J Anesth. 2008;22(3):248–52.

29. Hintong T, Chau-In W, Thienthong S, Nakcharoenwaree S. An analysis of the drug error problem in the Thai Anesthesia Incidents Study (THAI Study). Journal of the Medical Association of Thailand. 2005;88(Suppl 7):S118–27.

30. Webster CS, Merry AF, Larsson L, Mcgrath KA, Weller J. The Frequency and Nature of Drug Administration Error During Anaesthesia. Anaesth Intensive Care. 2001;29(5):494–500.

31. Fasting S, Gisvold SE. Adverse drug errors in anesthesia, and the impact of coloured syringe labels. Canadian Journal of Anesthesia. 2000;47(11):1060–7.

32. Charuluxananan S, Sriraj W, Lapisatepun W, Kusumaphanyo C, Ittichaikulthol W, Suratsunya T. Drug errors from the Thai anesthesia incidents monitoring study: Analysis of 1,996 incident reports. Asian Biomedicine. 2017;6(4):541–7.

33. Chopra V, Bovill JG, Spierdijk J. Accidents, near accidents and complications during anaesthesia. A retrospective analysis of a 10-year period in a teaching hospital. Anaesthesia. 1990;45(1):3–6.

34. Freestone L, Bolsin SN, Colson M, Patrick A, Creati B. Voluntary incident reporting by anaesthetic trainees in an Australian hospital. International Journal for Quality in Health Care. 2006;18(6):452–7.

35. Khan FA, Hoda MQ. Drug related critical incidents. Vol. 60, Anaesthesia. 2005. p. 48–52.

36. Labuschagne M, Robbetze W, Rozmiarek J, Strydom M, Wentzel M, Diedericks BJS, et al. Errors in drug administration by anaesthetists in public hospitals in the Free State. South African Medical Journal. 2011;101(5):324–7.

37. Maximous R, Wong J, Chung F, Abrishami A. Interventions to reduce medication errors in anesthesia: a systematic review. Vol. 68, Canadian Journal of Anesthesia. Springer; 2021. p. 880–93.

